# The effect of the foreign language on cognitive reappraisal during exposure to a phobic stimulus

**DOI:** 10.1101/2022.12.19.22283671

**Authors:** Isabel Ortigosa-Beltrán, Irene Jaén, Víctor Costumero, Azucena García-Palacios

## Abstract

The foreign language effect shows that emotional reactivity is reduced when we use a second language instead of our native one to address different situations. The present study aims to test whether the use of cognitive reappraisal could be influenced by the participant’s language (native/foreign). For this purpose, a sample of 60 participants with subclinical phobia to cockroaches was exposed to several phobic and neutral pictures while they used cognitive reappraisal in their native or in a foreign language. Physiological measures (pupil dilation and electrodermal activity) and self-reports of affective valence and arousal were collected. Results show an overall effectiveness of the strategy of reappraisal in both languages. Additionally, the use of a foreign language depicts a decrease in the affective negativity of the threat stimuli in terms of valence. The ratings of arousal also show a general higher arousal as an effect of the use of a foreign language. The present results suggest that using a foreign language could be advantageous to reduce negative emotionality by reappraisal. However, it could be a barrier for reappraising non-unpleasant pictures. Further studies should explore the foreign language effect in psychotherapy to open new ways of approaching different disorders.

## Introduction

The diversity of languages is growing. Bilingualism or multilingualism in a foreign country is becoming common as mobility between countries increases. Being in a foreign language context frequently implies the search for resources in a non-native language. These resources, among many others, may be the need to seek mental health professionals. In the same direction, mental health disorders are increasing as well, especially anxiety disorders. As such, it has become usual and sometimes inevitable to treat these disorders in the language of the host country. In a position in which someone needs to approach a phobia in a foreign language context, one could think that the treatment would not be as effective as if it were done in the native language. However, the use of a foreign language in areas such as decision-making (e.g., Cipolletti et al., 2016; Costa et al. 2014) and the acquisition of fear experiments (García-Palacios et al, 2018) have shown that the confrontation of negative emotional material in a second language reduces the emotional impact, and in consequence, it could facilitate the efficiency of some parts of the process.

The use of a non-native language to investigate affective processing has gained some focus in the past years (Pavlenko et al., 2012). Specifically, studies in areas such as decision making have pointed out how the use of a second language could affect processes involving moral choices differently (Costa et al., 2014) or the aversion to possible losses (Keysar et al., 2012). These studies found a pattern in which the decisions made in a foreign language resulted in more utilitarian and rational when confronting moral dilemmas, and the acceptance of gambles produced less aversion to losses, respectively. The second language had an impact on the results, causing a more deliberate and more impersonal reasoning and consequently more rational choices. This contrast under the influence of a non-native language was called the ‘foreign language effect’. Successive authors have discussed a plausible explanation for this phenomenon in emotional terms (Caldwell-Harris, 2015). Using a foreign language would imply a reduction in emotionality, evoking a psychological distance that would result in a ‘colder’ judgment compared to the familiarity of the native language. In this vein, a growing body of literature has focused on the influence of a second or foreign language on affective processing (Geipel et al., 2016; Hadjichristidis et al., 2019). For instance, some studies presenting emotional words or sentences in the native and in a foreign language showed reduced impact in the later one (Caldwell-Harris, 2015; Jończyk et al., 2016). Essentially, the scientific literature has addressed this effect on different levels, subjective, physiological and neural (e.g., Harris et al., 2015; Iacozza et al., 2017). Some examples directly relate the foreign language effect to a decrease in emotional reaction through physiological measures. Bilingual speakers showed reduced skin conductance responses (SCR) when listening to emotional phrases in their second language (English), compared to their native one (Turkish), (Caldwell-Harris & Aycicegi-Dinn, 2009). There is also a decrease in arousal when hearing swear words measured by skin conductance (Harris et al., 2003), also shown in pupil size results (Iacozza et al., 2017). Iacozza and colleagues presented highly emotional sentences to a group of participants in the native and in their second language, obtaining via a diminished pupil size response that reading the sentences aloud in the second language dampened the emotional load of the sentences. They suggested lighter emotional processing in a second language in the context of reading emotional material. According to research, what generates this differential effect in a foreign language and makes it distinctive from the native one, is the higher cognitive load associated with the foreign language processing (Caldwell-Harris & Ayçiçeği-Dinn, 2021). Specifically, it requires a greater lexical processing demand (Ivanova & Costa, 2008). In addition, Branzi et al., (2016) supported that using a foreign language leads a greater recruitment of neural areas involved with cognitive control.

The clinical setting has also shown a preference for a foreign language on some occasions, particularly when the content to be dealt with was emotionally charged (Dewaele, 2010; Marcos, 1976; Guttfreund, 1990). More importantly, clinical case studies with bilinguals provide evidence of psychotherapy in a foreign language being as effective as therapy in a native language (Griner & Smith, 2006). Also, in some cases a foreign language proved to be even more useful than native language to detach from traumatic childhood memories (Aragno & Schlachet, 1996). The intuition is that a foreign language can function as a protector, enabling patients to feel more distance when treating emotional experiences and, consequently, feel safer (Buxbaum, 1949; Movahedi, 1996).

More relevant to our research, one of the main strategies in the field of psychology which entail large linguistic charge, are emotion regulation strategies. They are commonly used in the treatment of emotional disorders, among others, and are often combined with other therapeutic strategies like exposure therapy to cope with a particular fear that the patient may have. Interestingly, the emotion regulation strategies models are based on language whether transform, reinterpret and regulate people’s emotions by verbally, mentally or writing procedures. These strategies contrast processes either implicitly (affect labeling) or explicitly (cognitive reappraisal) (Gyurak et al., 2011). Some studies provide evidence that explicit regulation strategies could approach the fearful object more efficiently, since the patient is aware of what he is doing (Beck et al., 2005) within the cognitive-behavioral therapy (CBT) in anxiety disorders. Koelsch et al., (2015) already posed that language can function as a form of emotional regulation through reappraisal. Particularly, the strategy of reappraisal refers to the transformation or reinterpretation of the situation in order to alter its emotional impact (Gross, 1998). Thus, the strategy of reappraisal implies robust evaluation of the thought patterns in order to reinterpret its meaning (Richards et al., 2003). In addition, this strategy involves language, usually in the form of inner speech (Salas et al., 2018). Hence reappraisal seems to provide a good approach to explore the mechanisms of the foreign language effect in the regulation of emotions. Flykt and Bjärtå (2008) pointed out that the additional controlled processing of higher resource demanding tasks modulates fear responses, and reduces arousal in physiological responses. In this line, literature seems to suggest that people using reappraisal report feeling significantly greater affective valence and less arousal when faced negative stimuli, indexed by diminished electrodermal activity and pupil dilation (e.g., Burklund, et al., 2014; Ray et al., 2010; Shahane et al., 2019).

Even though clinical records have traditionally shown some preferences for a second language in relation to emotional issues, there is still a lot to understand concerning the use of emotion regulation strategies, especially taking into account certain disparity in the functioning of the foreign language effect in the clinical context. Morawetz et al. (2017) supported the view of some emotion regulation techniques can benefit from the foreign language effect. They reported that content labeling was more effective in a second language, while reappraisal showed not being dependent on the language context. A more recent study by Vives et al. (2021) found in a neuroimaging study higher activation of the amygdala when using affect labeling in a foreign language, suggesting that a foreign language does not reduce emotionality with this strategy. These studies suggest that the foreign language effect could be an important factor in the regulation of emotions, yet it deserves further exploration.

With this background, certain doubts are raised so far in relation to the possible effect that the use of a foreign language can have on psychological strategies that deal with emotional issues. Previous studies of this line of research focused on the emotion of fear, and examined whether the foreign language effect influenced the acquisition and extinction of fear following an instructed fear paradigm in a series of experiments conducted by our team. The results concluded that the acquisition of fear was weaker in terms of physiological measures (skin conductance and pupil dilation) in a foreign language context (García-Palacios et al., 2018). However, this effect was not shown in the extinction process, resulting equally effective in both linguistic contexts (Ortigosa-Beltran et al., in press).

Due to the lack of research in the foreign language effect on psychotherapeutic processes as extinction and emotion regulation, here we aim to illuminate this area and go a step further. This study follows the line of a previous study where an extinction paradigm was used (Ortigosa-Beltran et al., in press), with the addition to a closer orientation to clinical practice. Specifically, the question it is attempted to address is whether using a different language modulates the efficacy of the strategy of cognitive reappraisal during a brief course of exposure to a fearful stimulus in people with subclinical phobia. We propose that the combination of effects of the strategy of reappraisal and the softener effect associated with the use of a foreign language help soften the levels of arousal when confronting fearful stimuli, as well as reduced unpleasant ratings. According to some examples in literature, we predict greater baseline arousal in the foreign language group in comparison with the native group due to the additional cognitive load (Alnaes et al., 2014). Additionally, we expect to find differences between regulation and non-regulation conditions. Specifically, we hypothesize that reappraisal will be associated with greater valence self-reports, as well as reduced arousal self-reports, and diminished physiological responses (pupil size and electrodermal activity), being these differences greater in the foreign group due to the distance associated with this language.

## Method

### Participants

Sixty participants (49 females, mean age = 22.31 years, SD = 2.73) were recruited from an initial sample of 248. The participants scored between the second and the third quartile on the Cockroaches Phobia Questionnaire (M = 67.01, SD = 31.22), adapted from the Spider Phobia Questionnaire (Klorman et al., 1974) in order to select the participants with innate rejection to the negative images presented. Participants had an intermediate/high level of English according to an adaptation from Marian et al., (2007). All participants gave informed consent and were compensated with six euros. The inclusion criteria were: Spanish as mother tongue; relatively proficient level of English measured with a self-perceived level of knowledge; less than one year living in an English-speaking country. The exclusion criteria were to have no psychiatric problem in immediate need of treatment; and no current alcohol or drug dependence. All participants had normal or corrected-to-normal vision and completed several questionnaires prior to the experiment. These questionnaires included a short sociodemographic questionnaire which examined their level of education and income, a questionnaire to explore anxiety trait and state (State-Trait Anxiety Inventory; STAI; Spielberger et al., 1983), a questionnaire for depression symptoms (BDI-II; Beck et al., 1996), and one for emotion regulation abilities (Emotion Regulation Questionnaire; ERQ; Gross & John, 2003). The participants were matched in age, education, income, anxiety levels, depression symptoms and emotion regulation strategies skills (see Table 1), and the number of participants per group and condition was similar (or above) to previous studies (García-Palacios et al. 2018).

**Table 1.** Participant’s characteristics in the native and in the foreign language groups (means and standard deviations).

|  | **Native (n = 30)** | **Foreign (n = 30)** |
| --- | --- | --- |
| Females (number) | 22 | 27 |
| Age (in years) | 23.31(2.9) | 21.95(2.8) |
| English level | 6.39(1.3) | 6.46(1.42) |
| Phobia | 66.86(29.8) | 69.78(35.5) |
| Money income (in euros) | 2187.50 | 2732.76 |
| STAI-E | 16.95(9.16) | 14.32(7) |
| STAI-R | 25.68(7.8) | 16.91(6.86) |
| BDI-II | 12.68(7.3) | 8.86(6.4) |
| ERQ | 38.77(6.4) | 37.23(9.8) |

Thirty participants were randomly assigned to the foreign language context (27 females) and 30 to the native language context (22 females). The mean level of English did not differ between the participants assigned to each group (see Table 2), nor did it differ between language skill types [Speaking: *t*(48) = -.25, *p* = .65; Listening: *t*(48) = -.32, *p* = .65; Writing: *t*(48) = -.62, *p* = .40; Reading: *t*(47) = -.07, *p* = .81]. Participants under 80% pupil validity (n=4) were excluded from the analysis, resulting in a group of 56 participants for the final analysis of pupil size, being 29 in the foreign language group and 27 in the native one. The sample resulted in 56 participants for pupil dilation analysis, 60 for electrodermal activity analyses and 59 for subjective ratings. The study was approved by the ethics committee at author’s university.

**Table 2.** Participant’s basic language skills and age of acquisition in the native and in the foreign language groups (means and standard deviations).

|  | **Native (n = 30)** | **Foreign (n = 30)** |
| --- | --- | --- |
| Speaking | 5.52(1.6) | 5.64(1.6) |
| Listening | 6.44(1.78) | 6.6(1.7) |
| Writing | 6(1.9) | 6.32(1.6) |
| Reading | 6.92(1.9) | 6.96(1.7) |
| Age of acquisition (years old) | 6.16(1.8) | 6.2(1.5) |

### Stimuli and design

Participants were randomly assigned to the language context prior to the beginning of the experiment. The task adapted the emotion regulation strategy of reappraisal in two language contexts during a brief course of exposure to negative and neutral pictures. A total of 40 trials were performed: 20 neutral (butterflies) and 20 negative (cockroaches). Pictures were chosen from the repository Grimaldos et al. (2021), using the normative values for affective valence and arousal in Spanish samples. Negative (valence: *M* = 2.01; *SD* = 0.96; arousal: *M* = 4.67; *SD* = 1.55); neutral (valence: *M* = 4.91; *SD* = 0.97; arousal: *M* = 3.93; *SD* = 1.51). These pictures were presented in a random order to both language context groups, equally associated with the conditions of ‘reappraisal’ and ‘nonregulation’. Right before the experiment started, a reminder with the instructions appeared on the screen in the corresponding language for 10 seconds. The trial structure was similar to previous literature in emotion regulation strategies and language (Langeslag & Van Strien, 2018; Morawetz et al. 2017). The beginning of each trial was a fixation cross for 10 seconds, 4 seconds of which served as a baseline for posterior analysis (see Figure 1). Following Bebko, Franconeri, Ochsner and Chiao (2011), the emotion regulation strategy cue was presented before the picture for 2 seconds, the cue ‘Look’ for the nonregulation items, and ‘Decrease’ for the reappraisal items, in the corresponding language. The instructions for the cue ‘Look’ were to simply view the picture on the screen without trying to avoid it or think of something else, only responding naturally to the stimulus, following previous studies (Fuentes-Sánchez et al. 2019; Jaen et al., 2021; Webb et al., 2012). The instructions for the trial cues with ‘Decrease’ were to keep in mind and say out loud the sentence ‘It can not do anything to me’ in the foreign language (English), and ‘No puede hacerme nada’ in the native language (Spanish), in order to reduce the intensity of the negative emotion. Generally, in monolingual studies using reappraisal the participants are trained to display a conscious and volitional strategy generating their own mental sentences to decrease the emotion (e. g. Daros et al., 2018; Fuentes-Sánchez et al., 2019). However, in this case we trained them to mentally go through the same reinterpretation, in both languages, with the purpose of having the same language content in both languages and to avoid the extra cognitive load in the foreign language group associated with the elaboration of a sentence in a non-native language. In addition, the reappraisal sentence was asked to be said out loud to avoid possible inner speech. Immediately after each cue, a picture stimulus was presented with a duration of 8 seconds. Afterwards, participants completed ratings of valence and arousal related to each picture according to the Self-Assessment Manikin (SAM; Lang, 1980) thus providing a measure of trial-by-trial emotion regulation success. In this nine-point scale the valence figures ranged between an unhappy figure to a smiling happy one, while the arousal dimension ranged from a relaxed figure to an agitated, excited one. The report was made by saying out loud in the corresponding language the degree of arousal of valence self-assessed by the participant. The trial ended with an interstimulus interval (ITI) of 10 seconds of duration.

**Figure 1.**
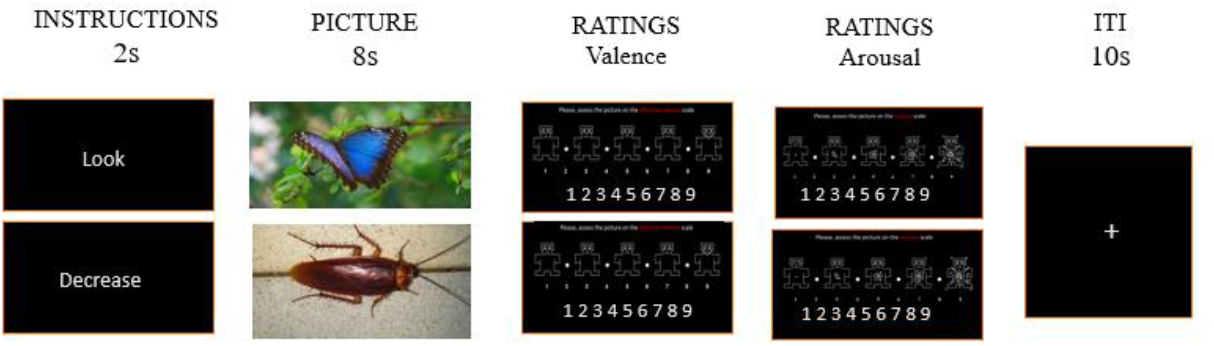
Trial design of the emotion regulation task.

### Psychophysiological data

The physiological measures recorded were the pupil size (Tobii Pro Lab) and the electrodermal activity (EDA; Shimmer3 GSR), both significant indicators of emotional charge in aversive stimuli. Pupil dilation has been shown to be a good measure of fear in the confrontation of fearful stimuli (Bradley et al., 2008; Hess & Polt, 1960), while the electrodermal activity is one of the main physiological measures in the automatic emotion responses (see Kreibig, 2010 for a review). The eye-tracker registered data at a sampling rate of 120 Hz. Images were displayed on a 19” monitor approximately 50 cm from the participants’ eyes. The screen monitored the pupil and served to display the task to the participants. Preprocessing for pupil dilation was carried out with MATLAB, following the guidelines provided by Kret and Sjak-Shie (2019). The raw data was filtered to remove invalid pupil size samples and artifacts and gaps in the sample were discarded, in order to obtain a smooth signal with valid data. Pupil size was averaged across both eyes and reduced to the mean value across the 8 second-trial. Electrodermal activity was recorded using a sampling rate of 125 Hz. Invalid data were removed in order to attain a continuous signal. Mean electrodermal activity was averaged for the 8-seconds trial. The electrodes were placed in the middle fingers of the non-dominant hand and remained steady for two minutes until the signal stabilized. The baseline for both physiological measures was calculated averaging the mean from the 4 seconds prior to each trial. The change scores were calculated as the difference between the mean of each trial in each condition and the mean of the baseline for both measures.

### Procedure

Participants completed online a self-perceived questionnaire related to their English level and a phobia to cockroaches questionnaire prior attending to the in person session, in order to see whether they fulfilled the inclusion criteria.

The session started with the participants reading the consent form and filling the questionnaires related to anxiety, depression, emotion regulation strategies and sociodemographic. Once the exclusion criteria were met, the participants were randomly assigned to one of the language conditions (native or foreign) and they were positioned in front of the computer screen where the experiment was carried out. The sensors of the electrodermal activity were placed in the middle and index fingers of the non-dominant hand remained there until the signal stabilized. Afterwards, the researcher explained the concept of emotion regulation strategies, focusing on the strategy of cognitive reappraisal, with examples. The language context began when the participant started the task. Firstly, a screen with the instructions indicated to the participant that a cue with the relevant word instruction, ‘Look’ or ‘Decrease’, would precede a picture. The instructions explained the task in each condition, with the support of the researcher reinforcing the written instructions. They were also informed in the instructions that a screen with self-reports of valence and arousal would be shown at the end of each trial, and they would have to report verbally in the corresponding language condition the self-perceived degree of arousal and valence. After assuring that the participant had understood the instructions, an example trial was shown to the participant to see the structure of the task. Right before starting the task, the eyes of the participant were calibrated on the screen with the eye-tracker. Once the electrodermal activity signal was stable and the eye-tracking calibrated, the task started. When the participant finished the 40 trials of the task, the sensors were removed and they completed the formulary to receive the payment.

The whole session was conducted in the corresponding language group, including all the interactions, verbal and written instructions and items, in order to provide language consistency. The session consisted of 10/15 minutes of questionnaires and 20/25 minutes of task in the computer, lasting around 40/45 minutes in total.

### Data analysis

Four separate 2 (Stimulus Type: negative vs neutral) x 2 (Regulation Strategy: reappraisal vs non-regulation) x 2 (Language Context: native vs foreign) repeated measures ANOVAs were performed for self-reported affective valence, self-reported arousal, pupil size, and electrodermal activity. Stimulus Type (negative vs neutral) and Regulation Strategy (Reappraisal vs non-regulation) were set as within-participant factors, and Language Context (native vs foreign) was set as a between-participants factor. Means and SDs are presented in Table 3. Assumptions of normality, homoscedasticity, sphericity, and equality of variances were explored using the Mauchly test and the Greenhouse-Geisser correction was used where appropriate. Additionally, post-hoc pairwise comparisons were performed using *t* tests to evaluate differences between stimuli types as well as between the reappraisal and non-regulation conditions when significant differences in main effects were found. Alpha level was set at 5% for the repeated measures ANOVAs and at 1% for *t* tests. Partial eta squared (*η*_*p*_^*2*^) and Cohen’s *d* were obtained as measures of effect size. All statistical tests were conducted using SPSS IBM Statistics version 23 and graphs were made with R (R Core Team, 2020).

**Table 3.** Means, standard deviations and confidence intervals by Language, Strategy and Condition.

|  | Native Language |  |  |  |  |  | Foreign Language |  |  |  |  |  |
| --- | --- | --- | --- | --- | --- | --- | --- | --- | --- | --- | --- | --- |
|  | Look |  |  | Reappraisal |  |  | Look |  |  | Reappraisal |  |  |
|  | 95% CI |  |  | 95% CI |  |  | 95% CI |  |  | 95% CI |  |  |
|  | Mean (SD) | Lower | Upper | Mean (SD) | Lower | Upper | Mean (SD) | Lower | Upper | Mean (SD) | Lower | Upper |
| **Valence self-reports** |  |  |  |  |  |  |  |  |  |  |  |  |
| Neutral | 6.94 (1.32) | 6.45 | 7.43 | 7.23 (1.16) | 6.75 | 7.71 | 7.04 (1.27) | 6.55 | 7.52 | 6.69 (1.37) | 6.21 | 7.17 |
| Negative | 2.93 (1.46) | 2.44 | 3.43 | 3.05 (1.23) | 2.62 | 3.48 | 2.36 (1.14) | 1.88 | 2.85 | 2.65 (1.02) | 2.23 | 3.07 |
| **Arousal self-reports** |  |  |  |  |  |  |  |  |  |  |  |  |
| Neutral | 2.09 (1.24) | 1.50 | 2.68 | 2.03 (1.10) | 1.4 | 2.5 | 2.50 (1.80) | 1.92 | 3.08 | 2.39 (1.70) | 1.85 | 2.92 |
| Negative | 4.76 (1.92) | 4.03 | 5.50 | 4.58 (1.77) | 3.8 | 5.2 | 6.10 (1.96) | 5.38 | 6.83 | 5.58 (1.90) | 4.90 | 6.27 |
| **Pupil size** |  |  |  |  |  |  |  |  |  |  |  |  |
| Neutral | -15.58 (5.98) | -17.61 | -13.55 | -15.84 (5.59) | -17.68 | -13.99 | -19.44 (4.20) | -21.44 | -17.45 | -14.10 (3.58) | -15.91 | -12.30 |
| Negative | -18.66 (6.23) | -20.78 | -16.54 | -16.90 (6.42) | -19.09 | -14.71 | -19.52 (4.40) | -21.60 | -17.44 | -17.65 (4.57) | -19.80 | -15.51 |
| **Electrodermal activity** |  |  |  |  |  |  |  |  |  |  |  |  |
| Neutral | -1.08 (1.32) | -1.74 | -0.42 | 0.28 (1.23) | -0.40 | 0.97 | -0.43 (2.16) | -1.12 | 0.24 | 0.63 (2.33) | -0.08 | 1.34 |
| Negative | -0.32 (1.68) | -0.91 | 0.263 | 2.00 (3.18) | 0.90 | 3.10 | 0.15 (1.47) | -0.46 | 0.76 | 1.73 (2.65) | 0.60 | 2.87 |

## Results

### Self-ratings

For valence self-reports, the main effect of Stimulus Type was significant [*F*(1, 55) = 63.99, *p* < .001, *η*_*p*_^*2*^= .54], but was not significant neither for Regulation Strategy (*F* < 1) nor Language Context [*F*(1, 55) = 2.26, *p* = .11, *η*_*p*_^*2*^= .08]. Specifically, valence was rated lower for negative than for neutral stimuli. The repeated measures ANOVA did not reveal differences between Stimuli Type x Language Context (*F* < 1), Regulation Strategy x Language Context [*F*(1, 55) = 2.13, *p* = .13, *η*_*p*_^*2*^= .07], nor Stimuli Type x Regulation Strategy (*F* < 1). However, Stimuli Type x Regulation Strategy x Language Context was significant [*F*(1, 55) = 5.29, *p* < .01, *η*_*p*_^*2*^= .16]. Specifically, post-hoc comparisons showed that participants in the foreign context rated negative stimuli with greater valence when they use reappraisal compared to non-regulation [*t*(29) = 2.43, *p* = .01, *d* = .27]. Also, they rated neutral stimuli with lower valence when they were instructed to reappraise compared to non-regulation [*t*(28) = 3.74, *p* < .001, *d* = 0.26]. Participants in the native context reported greater valence when reappraising neutral pictures compared to non-regulation [*t*(27) = 3.00, *p* < .01, *d* = .23], but no differences were found between reappraisal and non-regulation for negative pictures (*t* < 1) (see Figure 2 A).

**Figure 2.**
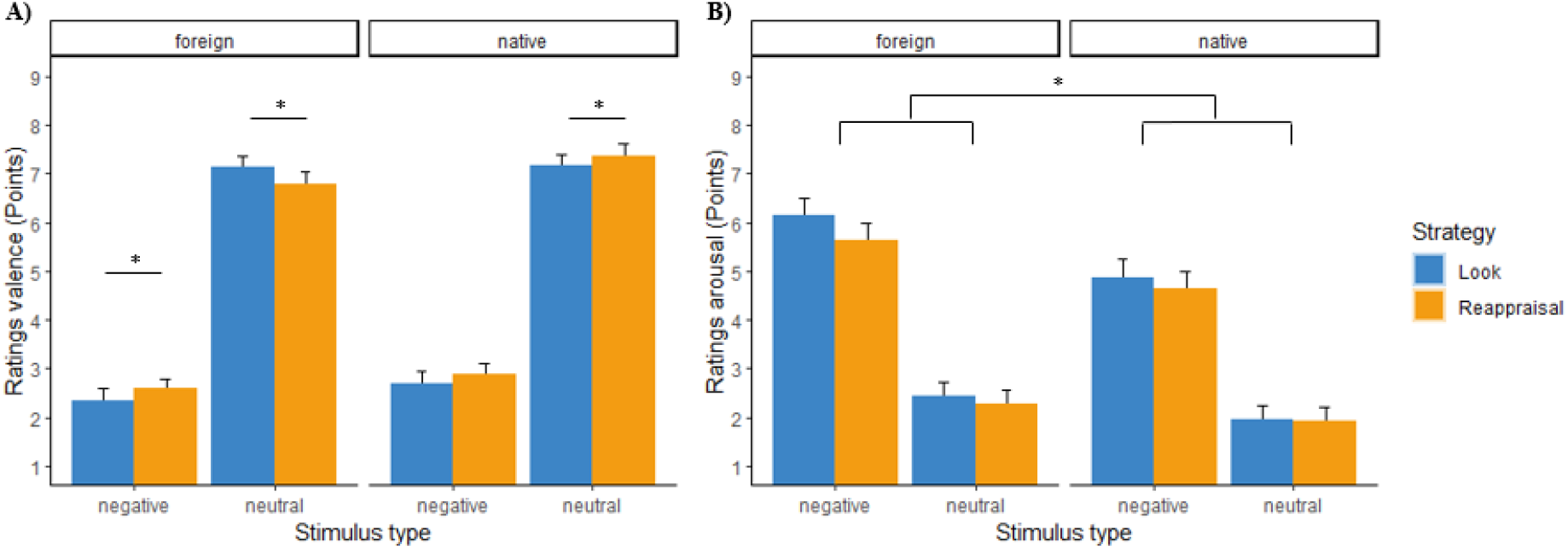
**(A)** Ratings of valence per Condition (neutral and negative) and Regulation Strategy (non-regulation in blue, reappraisal in orange), in each language group (foreign on the left, native on the right). The scale ranges from 0 (negative) to 9 (positive). **(B)** Ratings of arousal per Condition (neutral and negative) and Regulation Strategy (non-regulation in blue, reappraisal in orange), in each language group (foreign on the left, native on the right). The scale ranges from 0 (low arousal) to 9 (high arousal).

In terms of arousal self-reports, the main effect of Stimuli Type [*F*(1, 55) = 12.17, *p* < .001, *η*_*p*_^*2*^= 0.18] and Language Context [*F*(1, 55) = 3.69, *p* < .05, *η*_*p*_^*2*^= .12] were significant, while a main effect was not found for Regulation Strategy (*F* < 1). Specifically, negative stimuli were rated as more arousing than neutral stimuli. In addition, people in the foreign context rated images as more arousing than people in the native context. Interactions between Stimuli x Language (*F* < 1) and Strategy x Language [*F*(1, 55) = 1.5, *p* = .23, *η*_*p*_^*2*^= .05] were not significant, as well as the interaction Stimuli Type x Regulation Strategy x Language Context (*F* < 1) (see Figure 2 B).

### Electrodermal activity

The overall repeated measures ANOVA revealed a significant main effect of Stimuli Type [*F*(1, 54) = 18.32, *p* < .001, *η*_*p*_^*2*^= .25] and Regulation Strategy [*F*(1, 54) = 28.52, *p* < .001, *η*_*p*_^*2*^= .35], but did not reveal a significant main effect of Language Context (*F* < 1). Specifically, electrodermal activity responses were greater for the negative stimuli compared with the neutral stimuli. In addition, participants showed greater electrodermal activity responses when they used the reappraisal strategy compared to non-regulation (see Figure 3). Interactions between Stimuli Type x Strategy [*F*(1, 54) = 2.99, *p* = .08, *η*_*p*_^*2*^= .05], Stimuli Type x Language Context (*F* < 1), Strategy x Language Context (*F* < 1), and Stimuli x Strategy x Group (*F* < 1) were not significant.

**Figure 3.**
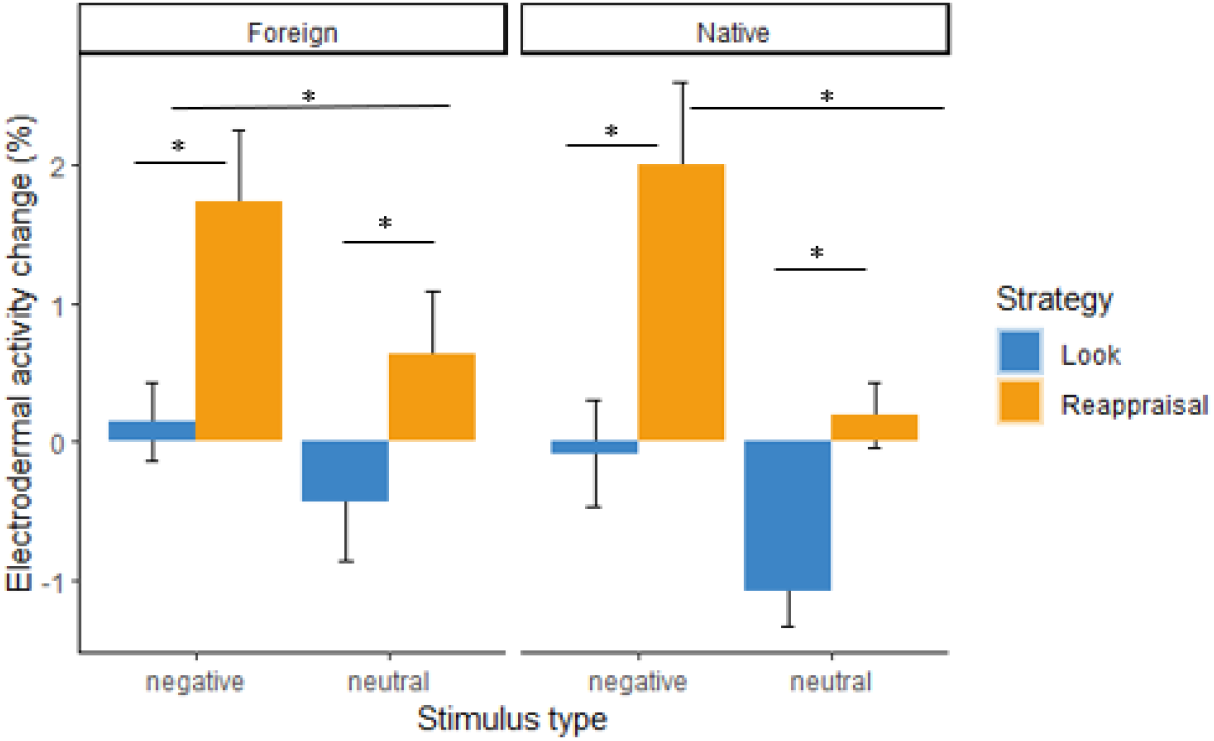
Electrodermal activity change in each language group with respect to the baseline epoch by Strategy and Condition. Graph on the left corresponds to the Foreign language context and graph on the right corresponds to the Native language context. Blue bars correspond to the non-regulation strategy and orange bars correspond to the reappraisal strategy. Error bars represent the standard error.

### Pupil size

A main effect was found for Stimuli Type [*F*(1, 51) = 31.98, *p* < .001, *η*_*p*_^*2*^= .39], and Regulation Strategy [*F*(1, 51) = 56.16, *p* < .001, *η*_*p*_^*2*^= .52]. However, the main effect of Language Context was not significant [*F* < 1]. Specifically, neutral stimuli produce greater pupil responses compared with negative stimuli. In addition, the reappraisal strategy was associated with greater pupil responses compared with the non-regulate condition. Regarding the interactions, Stimuli x Strategy [*F*(1, 56) = 3.56, *p* = .07, *η*_*p*_^*2*^= .07] and Stimuli Type x Language [*F* < 1] were not significant. However, the Strategy Type x Language Context interaction was significant [*F*(1, 51) = 24.06, *p* < .001, *η*_*p*_^*2*^= .32]. Specifically, post-hoc comparisons showed that effects of Strategy Regulation were not found for the native group [*t*(25) = 1.90, *p* = .04, *d =* .12], while pupil responses were higher for the foreign group when reappraisal strategy was used compared to non-regulation [*t*(26) = 8.50, *p* < .001, *d* = .89]. In addition, the Stimuli Type x Strategy x Language Context interaction was significant [*F*(1, 51) = 51.29, *p* < .001, *η*_*p*_^*2*^= .50]. As shown in Figure 4, the native group showed lower pupil responses during reappraisal compared to the non-regulation condition for the negative stimuli [*t*(25) = 3.6, *p* < .001, *d* = .28], but differences were not found for neutral stimuli (*t* < 1). The foreign group showed a significant effect of the Regulation Strategy for both negative [*t*(26) = 3.83, *p* < .001, *d* = .42] and neutral stimuli [*t*(26) = 11.16, *p* < .001, *d* = 1.37], being pupil responses lower when participants had to reappraise their emotions.

**Figure 4.**
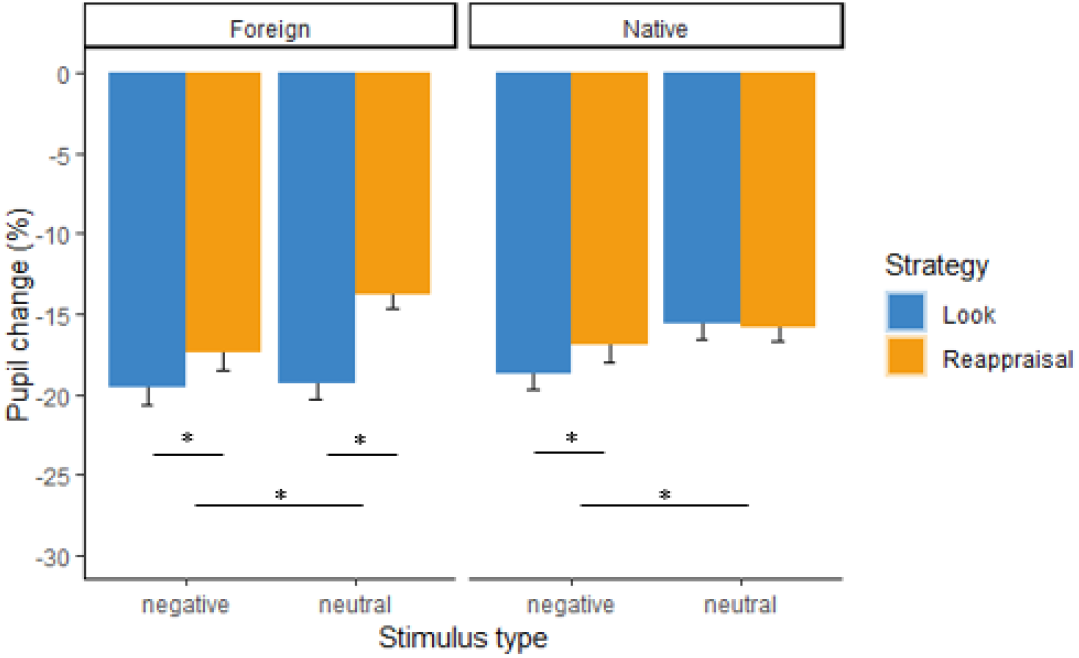
Pupil size change in each language group with respect to the baseline epoch by Strategy and Condition. Graph on the left corresponds to the Foreign language context and graph on the right corresponds to the Native language context. Blue bars correspond to the non-regulation strategy and orange bars correspond to the reappraisal strategy. Error bars represent the standard error.

## Discussion

This work follows a line of research that aims to explore the effect of the foreign language on processes involved in psychotherapy. Specifically, the purpose of the current study was to examine whether the use of a foreign language on bilingual people with subclinical phobia had an effect on the use of reappraisal during a short period of exposure to a fearful stimulus. Thus, a series of negative (cochraches) and neutral (butterflies) pictures were presented to participants with subclinical phobia to cockroaches while physiological and self-reported measures were collected. The task consisted of using either the reappraisal strategy or a non-regulation strategy in a random order, depending on the cue prior to each picture.

Overall, self-ratings of valence and arousal showed differences between pictures of cockroaches and butterflies. Similarly to previous studies using negative and neutral stimuli (Lang, Bradley & Cuthbert, 2008), negative pictures (i.e., cockroaches) were rated as less pleasant and more arousing than the neutral pictures (i.e., butterflies). In the same vein, electrodermal activity and pupil size response were greater during the visualization of cockroaches in comparison to butterflies, which is also associated with greater arousal during the visualization of negative pictures. For affective valence, these differences did not depend on the language context. For arousal, however, pictures of cockroaches were rated as more arousing, especially in the foreign language group. These results are in line with our prediction of a greater baseline arousal in the foreign language group in comparison with the native group. Based on previous literature, these differences are explained by the additional cognitive load (Alnaes et al., 2014). and higher anxiety associated with the use of a non-native language (MacIntyre et al., 1997) to regulate their emotions.

EDA also showed greater arousal when visualizing cockroaches, in contrast to pupil size response, which was higher during the trials in which butterflies were presented, compared with cockroaches. However, we believe that the significant main effect obtained for the stimuli type has been driven by the paradigm used here. This is, means for both stimuli categories could have been influenced by the interactions with the other factors included in the analyses. Also, it is possible that these results were produced by the difference of colors and luminance used in each picture category (Kohn & Clynes, 1969), which were not controlled in this study. Future studies should be conducted using grey images instead of colored ones.

Regarding the emotion regulation effects, valence self-ratings showed that using a foreign language is more effective than the native one to reappraise negative emotions. In particular, the foreign language group showed greater valence self-ratings during the reappraisal of negative pictures compared with non-regulation. However, the native context was less effective down-regulating their negative emotions, revealing no significant differences between reappraisal and non-regulation conditions. These findings are in line with our hypothesis, suggesting that foreign language can be used to attenuate negative emotions when confronting fearful stimuli. Interestingly, these findings were not found for neutral pictures, in which participants rated as more negative the butterflies when reappraising. It may indicate that the suggested detachment produced by the use of a foreign language (García-Palacios et al., 2018) can be useful for reappraising negative stimuli. However, it could hamper the use of reappraisal for pictures that are not unpleasant for the participant. It is worthy to note that, in this study valence and arousal self-ratings for butterflies were more positive (valence: M = 7.1, SD = 1.30: arousal: M = 2.24, SD = 1.49) than neutral, as previous studies had stated (Grimaldos et al., 2021; valence: M = 4.92, SD = 1.30; arousal: M = 3.93, SD = 1.51). This could be related to the comparisons carried out during the repository validation and the specific contrast here, only with cockroaches. Therefore, on the basis of the present results, we argue that the use of reappraisal using a foreign language can be advantageous to help reducing the perception of negative emotions, since the cognitive load associated with the use of a less familiar language helps to reduce emotionality. However, it is recommended to be cautious with the generalization of this effect, since it seems that the reduction of emotionality could be a barrier in the case of using reappraisal to increase positive emotions.

Psychophysiological measures, however, did not shown the expected results. The present study found that both, electrodermal and pupillary responses, were greater when participants had to reappraise their emotions compared with non-regulating, indicating an increasing of arousal. These results are in line with the pattern observed in previous studies, in which the use of reappraisal was associated with higher arousal in comparison to non-regulation trials (Bernat et al., 2011; Fuentes-Sánchez et al., 2019; Jaén et al., 2021), which has been commonly explained by the cognitive effort made to regulate emotions. Following that line, the physiological results obtained in this study are explained in terms of an increase of arousal produced by the cognitive load associated with the regulation of emotions. Of note, this increment in arousal during reappraisal might only indicates the cognitive effort implied in the process of regulation, but not the effectivity in this regulation.

With regard to the effects of the foreign language on the physiological measures, these were only found for pupil size. Specifically, participants in the foreign language group showed emotion regulation effects for both negative and neutral pictures. However, the native language group showed emotion regulation differences only for negative pictures, but not for neutral ones. Based on the studies that stated pupil dilatation as a marker of both cognitive effort and emotional processing in relation to emotion regulation strategies (Kinner et al., 2017), we state that our results support an effect when using a foreign language. Particularly, these findings support the idea of the additional cognitive load (Alnaes et al., 2014) associated with the use of a non-native language when participants are instructed to regulate their emotions, especially when participants are reappraising positive ones. Nevertheless, it is worth noting that the reappraisal strategies used in this study were focused on down-regulate negative emotions. Future studies should be carried out to determine the foreign language effect with more appropriate strategies for reappraising positive pictures.

These results contrast with the findings obtained by Morawetz et al. (2017), who reported that content labeling was more effective in a second language, while reappraisal showed not being dependent on the language context. However, there are some differences between the study conducted by Morawetz et al. (2017) and this study that can be highlighted. First, the participants were healthy in the Morawetz et al. (2017), whereas in our study a selection of participants with moderate scores on the Cockroaches Phobia Questionnaire were recruited. Moreover, participants in the study of Morawetz et al. (2017) used their inner speech during cognitive reappraisal, while in our study we instructed participants to say aloud the reappraisal strategy. The variances between the two studies could involve a more effectivity in the use of reappraisal in our study, explaining the discrepancies between them.

Furthermore, Vives et al. (2021) found that downregulation through affect labeling was not effective in the foreign language as compared to the native one. In fact, amygdala activation increased in the foreign language condition, suggesting that the cognitive load associated with the foreign language could interfere with an appropriate downregulation. In this study, however, we also obtained increased electrodermal and pupil responses associated with the cognitive load, but the findings obtained by self-reports indicated that the reappraisal strategy was effective in terms of increasing hedonic valence.

This study expands the current knowledge on the boundaries of the foreign language effect. Previous studies in this line of research showed a weakener effect of the conditioning during the acquisition of fear produced by the foreign language effect (Garcia-Palacios et al., 2018), as the participants put distance with the learning of the association with a possible aversive stimulus. However, this detachment effects were found in the process of extinction when the instructions changed in order to create the new learning of safety (Ortigosa-Beltran et al., in press). In this study we go one step further, introducing reappraisal verbal instructions during the exposure to cockroaches’ pictures to participants with subclinical phobia. The findings obtained have important clinical implications, suggesting that foreign language could be used in psychotherapy as a way to reduce emotionality during exposure sessions for specific phobia, in which psychotherapists want to achieve a decrease of fear when confronting threatening stimuli. In addition, this study opens the door to future studies that aim to study the effects of using a foreign language during exposition for the treatment of other pathologies such as prolonged grief disorder or post-traumatic stress disorder.

This study has some limitations that further studies should consider. In order to provide a more adequate context for the foreign language effect to be present, it would be recommended to provide a design with longer language involvement. Here we eliminated the possible inner speech to ensure that differences could not be explained by changes in the verbal content. However, this measure precludes the spontaneity of the strategy, which could reduce their effectivity. It would be recommendable to enhance spontaneity in order to increase the general language engagement in each language group. In addition, our study was conducted with participants with subclinical phobia. Further studies should explore the implications of this effect on a clinical group in order to test whether the presence of the foreign language effect could depend on the degree of anxiety or fear related to a disorder. Another limitation is the use of dark inter-stimulus screens, instead of the usual gray or white screens in the design of the study. Although it did not interfere with the interpretation of the results, it would be recommended to plan following studies with the usual clearer screens to avoid the possible influence of brightness on the modulation of autonomic nervous system responses (Vasquez-Rosati er al., 2017). Also, because only 12% of participants were male, an important question would be whether the current findings should be viewed as limited to females. We also suggest a deeper exploration of other types of bilingualism, varying characteristics such the level of proficiency or the context of acquisition. Future work with emotion regulation strategies and other paradigms could examine how different groups of bilinguals may show different patterns of results. These results cannot be generalized to other characteristics.

To conclude, the present study is the first empirical experiment on the influence of the foreign language effect on reappraisal in patients with subclinical specific phobia, showing the possible advantages to use it in the context of exposition to aversive stimuli. Thus, it contributes to the understanding of the role of foreign language in emotion regulation paradigms, an area that lacks of research. Future studies are needed to shape new ways of approaching to different disorders such as specific phobias within the field of psychotherapy.

## Data Availability

All data produced are available online at Open Science Framework (OSF)

https://osf.io/tjx3d/

## Competing interests

The authors declare that this study was carried out in the absence of any personal, professional or financial relationship that could be interpreted as a conflict of interest.

## Acknowledgments

The authors would like to thank the MultiMind project for its support in every way and for providing all the facilities.

## Availability of materials

The datasets analyzed for this study can be found in the Open Science Framework (OSF) Repository [https://osf.io/tjx3d/].

## Notes

**Funding** This work was supported by Marie Sklodowska-Curie Innovative Training Networks (ITN-ETN)—Multimind (H2020 MSCA-ITN-2017, grant 765556).

### Competing Interest Statement

The authors have declared no competing interest.

### Funding Statement

This work was funded by Marie Sklodowska-Curie Innovative Training Networks (ITN-ETN) Multimind (H2020 MSCA-ITN-2017, grant 765556).

### Author Declarations

Ethics committee of Universitat Jaume I de Castellon de la Plana gave ethical approval for this work

